# Influence of COVID-19 confinement measures on appendectomies in Germany – administrative claims data analysis of 9,797 patients

**DOI:** 10.1101/2020.09.25.20198986

**Authors:** Matthias Maneck, Christian Günster, Hans-Joachim Meyer, Claus-Dieter Heidecke, Udo Rolle

## Abstract

**Purpose:** COVID 19 pandemic had multiple influences on the social, industrial and medical situation in all affected countries. Measures of obligatory medical confinement were suspensions of scheduled non-emergent surgical procedures and outpatients’ clinics as well as overall access restrictions to hospitals and medical practices.

The aim of this retrospective study was to assess if the obligatory confinement (lockdown) had an effect on the number of appendectomies (during and after the period of lockdown).

**Methods:** This retrospective study was based on anonymized nationwide administrative claims data of the German Local General Sickness Fund (AOK). Patients admitted for disease of the appendix (ICD-10: K35-K38) or abdominal and pelvic pain (ICD-10: R10) who undergone an appendectomy (OPS: 5-470) were included. The study period included six weeks of German lockdown (16.03.-26.04.2020) as well as six weeks before (03.02.-15.03.2020) and after (27.04.-07.06.2020). These periods were compared to the respective in 2019.

**Results:** Overall number of appendectomies was significantly reduced during the lockdown time in 2020 compared to 2019. This decrease affects only appendectomies due to acute simple and non-acute appendicitis. Numbers for appendectomies in acute complex appendicitis remained unchanged. Female patients and in the age group 1-18 years showed the strongest decrease in number of cases.

**Conclusion:** The lockdown in Germany resulted in a decreased number of appendectomies. This affected mainly appendectomies in simple acute and non-acute appendicitis, but not complicated acute appendicitis. The study gives no evidence, that the confinement measures resulted in a deterioration of medical care for appendicitis.

**Authors contributions:** MM performed study conception, acquisition of data, analysis and interpretation of data, drafting the manuscript. CG performed study conception, analysis and interpretation of data, drafting the manuscript, critical revision of manuscript.

HJM performed study conception and critical revision of manuscript.

CDH performed study conception and critical revision of manuscript.

UR performed study conception, drafting the manuscript and critical revision of manuscript

## Introduction

COVID-19 pandemic had multiple influences on the social, industrial and medical situation in the affected countries. Confinement measures to minimize the number of infected persons included social distancing, avoidance of contact and formal lockdown in respective regions.

Medical confinement measures had been introduced from 16 March 2020 in Germany, with suspension of all scheduled hospitalizations, elective operations, outpatients’ clinics, stoppage of screening measures (e.g. mammography) and reduced opening hours of practices. Main aspect of all measures was to avoid contacts in the medical setting and spare protective equipment.

It had been assumed, that COVID-19 pandemic would not only have had a major impact on the delivery of elective care but also on emergency procedures. A recent report from Italy has shown a reduced rate of hospital admissions for acute coronary syndrome during COVID-19 outbreak [1].

Appendectomy is one of the most frequent abdominal surgeries in all age groups. Appendectomy usually would be performed in patients with simple acute and complex acute appendicitis.

Early reports showed increased number of complicated appendicitis during confinement period in GB [2]. This has also been shown in a small case series, which presented 7 pediatric cases with advanced appendicitis due to delayed presentation [3].

Medical associations in Germany expressed concerns that the medical confinement measures might lead an increase of patients with complicated appendicitis due to the delay of presentation to the hospitals ([4].

The aim of this retrospective study was to assess if the medical lockdown in Germany had an effect on the number of appendectomies (during and after the period of lockdown).

## Material and methods

This retrospective cohort study was based on anonymized nationwide administrative claims data of the German Local General Sickness Fund (AOK), the largest provider of statutory health care insurance in Germany. The AOK covers approximately 30% of the population. The claims data includes age, sex as well as data from inpatient episodes, including diagnoses, procedures and length of stay. Diagnoses were coded according to the 10th revision of the International Classification of Diseases (ICD-10). Procedures were documented using the German version of the International Classification of Procedures in Medicine (ICPM), the “Operationen-und Prozedurenschlüssel” (OPS).

We included patients aged ≥ 1 year admitted for disease of the appendix (ICD-10: K35-K38) or abdominal and pelvic pain (ICD-10: R10) who have undergone an appendectomy (OPS: 5-470). The study period included the six weeks of German lockdown (16.03.-26.04.2020) as well as six weeks before (03.02.-15.03.2020) and after (27.04.-07.06.2020). These three periods correspond to the calendar weeks: 6-11 (before), 12-17 (lockdown), 18-23 (easing). Similarly, patients were included for 2018 and 2019 with respect to the Easter holiday in 2020 (calendar week 15). To avoid confounding effects, the calendar weeks of 2018 and 2019 were shifted by +2 and -1 in respect to the Easter holidays respectively.

The primary outcome was incidence of hospital admissions. We calculated incidence rates for the primary outcome by dividing the number of cumulative admissions by the number of days for each time period.

Patients were stratified by appendicitis stage, gender and age. Appendicitis stages were classified as complex acute appendicitis (CAA), simple acute appendicitis (SAA) and non-acute appendicitis (NAA). CAA exhibit a generalized or localized peritonitis with perforation or rupture or a peritoneal abscess (ICD-10: K35.2, K35.31, K35.32). SAA was acute but without the aforementioned properties (ICD-10: K35.30, K35.8). NAA included other or non-specified types of appendicitis, other diseases of the appendix or abdominal and pelvic pain (ICD-10: K36, K37, K38, R10). According to their age patients were divided into three groups: 1-18 years, 19-64 years and ≥65 years.

Patient demographics, appendicitis stage and length of stay were summarized as descriptive statistics. Categorical data were presented as percentage, numeric data as mean with standard deviation, respectively. Trends among the three periods within 2020 were compared using univariate χ^2^ or Kruskal-Wallis tests using a significance level of 0.05.

Case reductions were determined by incidence-rate ratios (IRR) comparing each period of 2020 with the corresponding calendar week of 2019 using Poisson regression to model the number of admissions per day [1]. Bonferroni adjustment was done to correct P-values for comparing multiple patient groups (factor 144).

To investigate annual changes in the number of cases unrelated to the pandemic periods of 2019 were additionally compared to 2018.

All evaluations were performed with the software STATA16.0 (StataCorp, College Station, Texas).

## Results

### Appendectomy rates in 2020

The study comprised 9,797 AOK cases who undergone an appendectomy in 2020. Mean age was 34.5 years and 48.0% were female patients. Overall 23.7% were diagnosed for CAA, 70.1% for SAA and 6.2% for NAA. Detailed descriptive statistics for each period during the pandemic were shown in table 1. With respect to all patients the periods before, during and after the lockdown significantly differ in mean incidence per day, age, proportion of female sex, CAA and NAA.

**Table 1:** Patient Demographics of hospital admissions with appendectomy in 2020 (ALL: all appendicitis stages; CAA: complex acute appendicitis; SAA: simple acute appendicitis; NAA: non acute appendicitis).

| **Week number** | **Before**<br>**6-11** | **Lockdown**<br>**12-17** | **Easing**<br>**18-23** | **P** |
| --- | --- | --- | --- | --- |
| **ALL Cases** | 3,591 | 2,914 | 3,292 |  |
| Incidence rate per day | 85.5 | 69.4 | 78.4 | **<0.001** |
| Age in years |  |  |  |  |
| Mean (sd) | 32.7 (19.0) | 34.5 (19.8) | 34.9 (19.9) | **<0.001** |
| 1-18 (%) | 26.6 | 24.0 | 23.1 | **0.002** |
| 19-64 (%) | 66.0 | 67.1 | 67.2 | 0.527 |
| ≥ 65 (%) | 7.4 | 9.0 | 9.8 | **0.002** |
| Female sex (%) | 49.1 | 46.1 | 48.5 | **0.042** |
| Appendicitis (%) |  |  |  |  |
| Complex acute | 21.2 | 27.0 | 23.6 | **<0.001** |
| Simple acute | 70.1 | 69.2 | 70.8 | 0.390 |
| Non acute | 8.6 | 3.8 | 5.7 | **<0.001** |
| LOS, d; mean (sd) | 4.3 (3.6) | 4.3 (3.6) | 4.3 (3.2) | 0.567 |
| **CAA Cases** | 763 | 786 | 776 |  |
| Incidence rate per day | 18.2 | 18.7 | 18.5 | 0.8373 |
| Age in years |  |  |  |  |
| Mean (sd) | 42.4 (23.1) | 41.9 (23.2) | 44.2 (23.5) | 0.118 |
| 1-18 (%) | 21.9 | 21.4 | 19.1 | 0.348 |
| 19-64 (%) | 59.4 | 59.9 | 59.0 | 0.935 |
| ≥ 65 (%) | 18.7 | 18.7 | 21.9 | 0.191 |
| Female sex (%) | 41.0 | 41.2 | 43.3 | 0.603 |
| LOS, d; mean (sd) | 6.9 (5.2) | 6.8 (5.0) | 6.9 (4.7) | 0.844 |
| **SAA Cases** | 2,518 | 2,016 | 2,330 |  |
| Incidence rate per day | 60.0 | 48.0 | 55.5 | **<0.001** |
| Age in years |  |  |  |  |
| Mean (sd) | 30.1 (17.0) | 31.8 (17.7) | 32.2 (17.8) | **<0.001** |
| 1-18 (%) | 27.7 | 25.2 | 24.4 | **0.026** |
| 19-64 (%) | 67.6 | 69.1 | 69.4 | 0.371 |
| ≥ 65 (%) | 4.7 | 5.6 | 6.2 | 0.061 |
| Female sex (%) | 49.0 | 46.9 | 49.0 | 0.273 |
| LOS, d; mean (sd) | 3.6 (2.5) | 3.4 (2.1) | 3.5 (1.8) | **<0.001** |
| **NAA Cases** | 310 | 112 | 186 |  |
| Incidence rate per day | 7.4 | 2.7 | 4.4 | **<0.001** |
| Age in years |  |  |  |  |
| Mean (sd) | 29.1 (15.4) | 31.5 (15.1) | 30.2 (15.9) | 0.186 |
| 1-18 (%) | 29.0 | 19.6 | 22.6 | 0.086 |
| 19-64 (%) | 69.0 | 79.5 | 73.7 | 0.095 |
| ≥ 65 (%) | 1.9 | 0.9 | 3.8 | 0.230 |
| Female sex (%) | 70.0 | 66.1 | 62.9 | 0.257 |
| LOS, d; mean (sd) | 3.7 (2.4) | 4.1 (4.4) | 4.1 (3.6) | 0.577 |

LOS: length of stay

With beginning of the lockdown the daily case rate fell and increased again as the relaxations took effect, but not to previous levels (85.5, 69.4, 78.4, p<0.001). This trend was also observed when considering only SAA or NAA cases. However, for CAA cases no significant differences between the periods were observed.

These appendicitis-stage specific changes in the daily case rate result in different distributions of appendicitis-stages in the three periods. Within the lockdown and easing period, the proportion of CAA cases is higher and the proportion of NAA cases lower as compared to the before period. The difference in the easing period is not as strong as during the lockdown (CAA: 21.2%, 27.0%, 23.6%, p<0.001; NAA: 8.6%, 3.8%, 5.7%, p<0.001). Furthermore, the patient age and gender were influenced by lockdown and easing. While the mean age significantly increased during lockdown and easing (32.7 vs. 34.5 and 34.9 years; p<0.001), the proportion of women decreased particularly during lockdown (49.1% vs. 46.1% vs. 48.5%, p=0.042).

A significant change in length of stay was only observed for SAA cases (3.6 vs. 3.4 vs. 3.5, P<0.001). It was shortened during the lockdown and increased again after the easing, but not to the old level.

### Comparison of appendectomy rates 2020 to 2019 and 2019 to 2018

The examination of the three periods within 2020 already showed some effects of the lockdown and the easing on the study population. In the following, the three periods were examined within subgroups in relation to a common reference point, the previous year (2019).

Figure 1 shows the weekly numbers of cases for 2018, 2019 and 2020. Overall, as well as for SAA and NAA cases the weekly case number decreases with begin of the lockdown. In the easing period the weekly case number increases again, but not to the pre-lockdown level. For CAA cases no effects were visible.

**Figure 1:**
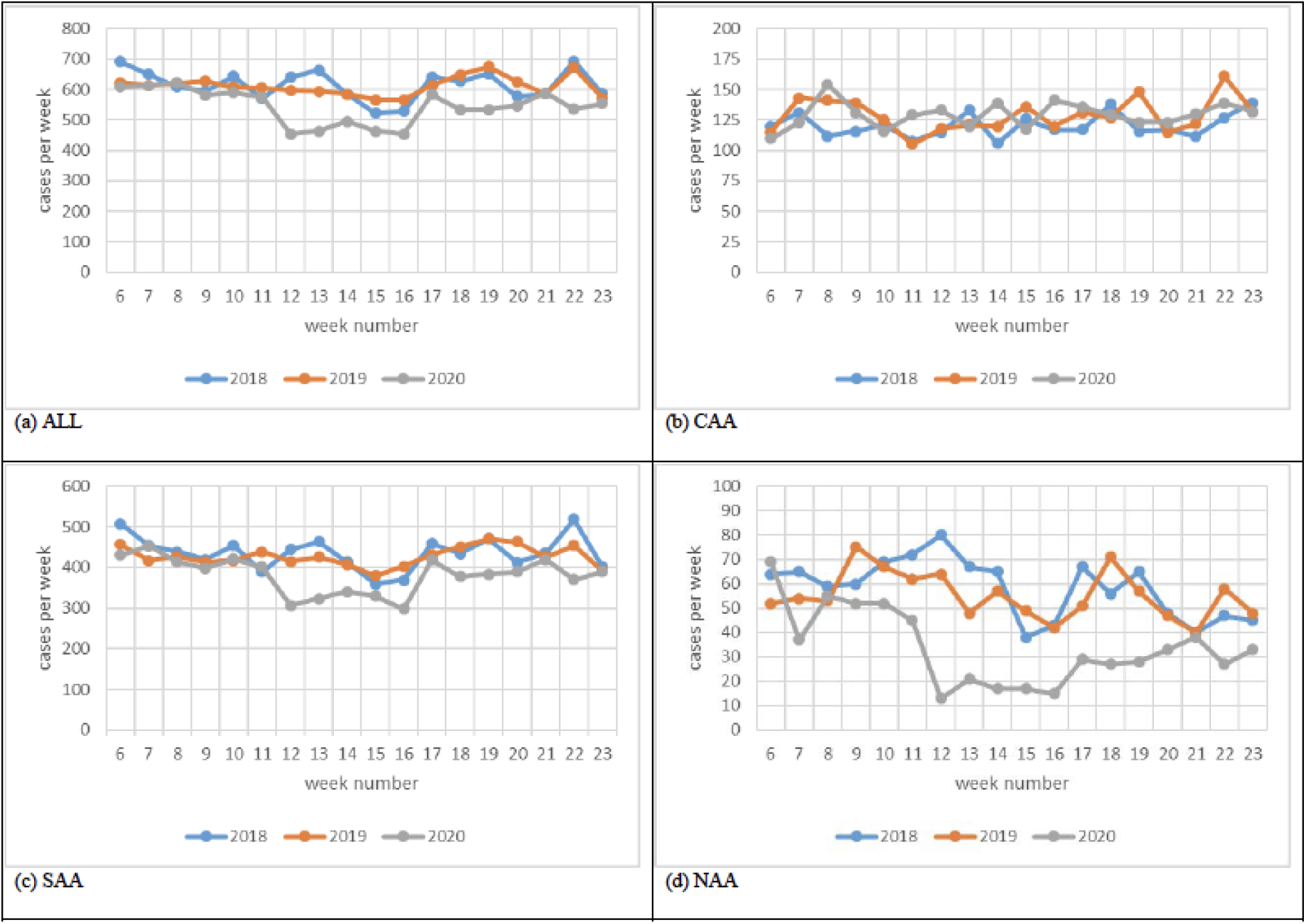
Weekly case of patients with appendectomy. (a) ALL: all appendicitis stages, (b) CAA: complex acute appendicitis, (c) SAA: simple acute appendicitis, (d) NAA: non-acute appendicitis.

Looking at all cases, a significant case reduction was observed for the lockdown (IR: 0.83, p<0.001) and easing period (IR: 0.87, p<0.001). However, only patients with SAA (lockdown IR: 0.82, p<0.001; easing IR: 0.88, p=0.001) and especially NAA (lockdown IR: 0.32, p<0.001; easing IR: 0.58, p<0.001) were affected.

Regarding age groups, there was a significant case reduction in patients aged 1-18 and 19-64 in both the lockdown (1-18 IRR: 0.74, p<0.001; 19-64 IRR: 0.85, p<0.001) and the easing period (1-18 IRR: 0.76, p<0.001; 19-64 IRR: 0.89, p=0.021). Patients aged 1-18 were most affected.

Considering gender, women in lockdown (IR: 0.79, p<0.001) and easing (IR: 0.84, p<0.001) and men only in lockdown period (IR: 0.86, p=0.002) showed a significant case reduction. The reduction was stronger for women.

When viewed together, gender and age-specific effects were observed even if only SAA or NAA were considered. Particularly strong case reductions were observed in women aged 1-18 with SAA in both lockdown (IR: 0.67, p<0.001) and easing (IR: 0.73, p=0.011). The same applies to the NAA cases, only that the decrease in the number of cases is even stronger (lockdown IR: 0.23, p<0.001; easing IR: 0.42, p<0.015).

For CAA cases as well as patients aged ≥65 no significant effects were observed.

To examine yearly case reductions without the influence of the pandemic all comparisons shown in table 2 were also done comparing 2019 to 2018. No significant case reductions were observed.

**Table 2:** Incidence rate ratios (IRR) and adjusted p-values of hospital admissions with appendectomy in 2020 in reference to 2019 estimated by Poisson regression. ALL – all appendicitis stages; CAA – complex acute appendicitis, SAA simple acute appendicitis, NAA – non-acute appendicitis.

| Gender,<br>age | ALL |  |  | CAA |  |  | SAA |  |  | NAA |  |  |
| --- | --- | --- | --- | --- | --- | --- | --- | --- | --- | --- | --- | --- |
|  | before | lockdow | easing | before | lockdow | easing | before | lockdow | easing | before | lockdow | easing |
| All cases | 0.97<br>1.000 | **0.83**<br>**<0.001** | **0.87**<br>**<0.001** | 0.99<br>1.000 | 1.05<br>1.000 | 0.96<br>1.000 | 0.98<br>1.000 | **0.82**<br>**<0.001** | **0.88**<br>**0.001** | 0.85<br>1.000 | **0.36**<br>**<0.001** | **0.58**<br>**<0.001** |
| 1-18 | 0.91<br>1.000 | **0.74**<br>**<0.001** | **0.76**<br>**<0.001** | 1.00<br>1.000 | 0.99<br>1.000 | 0.87<br>1.000 | 0.91<br>1.000 | **0.73**<br>**<0.001** | **0.77**<br>**0.001** | 0.80<br>1.000 | **0.28**<br>**<0.001** | **0.46**<br>**0.004** |
| 19-64 | 1.01<br>1.000 | **0.85**<br>**<0.001** | **0.89**<br>**0.021** | 1.04<br>1.000 | 1.09<br>1.000 | 0.98<br>1.000 | 1.01<br>1.000 | **0.85**<br>**0.001** | 0.9<br>0.279 | 0.90<br>1.000 | **0.41.00**<br>**<0.001** | **0.66**<br>**0.022** |
| ≥65 | 0.90<br>1.000 | 0.92<br>1.000 | 1.03<br>1.000 | 0.87<br>1.000 | 1.01<br>1.000 | 1.01<br>1.000 | 1.00<br>1.000 | 0.92<br>1.000 | 1.16<br>1.000 | 0.46<br>1.000 | 0.07<br>1.000 | 0.33<br>1.000 |
| Women | 0.94<br>1.000 | **0.79**<br>**<0.001** | **0.84**<br>**<0.001** | 0.98<br>1.000 | 1.07<br>1.000 | 0.93<br>1.000 | 0.96<br>1.000 | **0.81**<br>**<0.001** | 0.87<br>0.065 | 0.81<br>1.000 | **0.33**<br>**<0.001** | **0.53**<br>**<0.001** |
| 1-18 | 0.90<br>1.000 | **0.64**<br>**<0.001** | **0.67**<br>**<0.001** | 0.97<br>1.000 | 0.90<br>1.000 | 0.58<br>0.364 | 0.88<br>1.000 | **0.67**<br>**<0.001** | **0.73**<br>**0.011** | 0.88<br>1.000 | **0.23**<br>**<0.001** | **0.42**<br>**0.015** |
| 19-64 | 0.96<br>1.000 | **0.85**<br>**0.048** | 0.89<br>0.591 | 1.04<br>1.000 | 1.25<br>1.000 | 1.02<br>1.000 | 0.98<br>1.000 | 0.86<br>0.807 | 0.9<br>1.000 | 0.79<br>1.000 | **0.38**<br>**<0.001** | **0.61**<br>**0.035** |
| ≥65 | 0.9<br>1.000 | 0.83<br>1.000 | 1.07<br>1.000 | 0.84<br>1.000 | 0.83<br>1.000 | 1.07<br>1.000 | 1.05<br>1.000 | 0.90<br>1.000 | 1.19<br>1.000 | 0.43<br>1.000 | 0.00<br>1.000 | 0.27<br>1.000 |
| Men | 1.00<br>1.000 | **0.86**<br>**0.002** | 0.90<br>0.227 | 1.01<br>1.000 | 1.05<br>1.000 | 0.99<br>1.000 | 1.00<br>1.000 | **0.83**<br>**0.001** | 0.89<br>0.341 | 0.97<br>1.000 | **0.43**<br>**0.002** | 0.68<br>1.000 |
| 1-18 | 0.93<br>1.000 | 0.83<br>0.759 | 0.87<br>1.000 | 1.02<br>1.000 | 1.06<br>1.000 | 1.14<br>1.000 | 0.93<br>1.000 | 0.78<br>0.279 | 0.82<br>1.000 | 0.54<br>1.000 | 0.44<br>1.000 | 0.56<br>1.000 |
| 19-64 | 1.05<br>1.000 | **0.86**<br>**0.039** | 0.90<br>1.000 | 1.04<br>1.000 | 1.00<br>1.000 | 0.96<br>1.000 | 1.04<br>1.000 | 0.84<br>0.067 | 0.90<br>1.000 | 1.19<br>1.000 | 0.48<br>0.139 | 0.77<br>1.000 |
| ≥65 | 0.90<br>1.000 | 1.01<br>1.000 | 0.98<br>1.000 | 0.89<br>1.000 | 1.20<br>1.000 | 0.95<br>1.000 | 0.95<br>1.000 | 0.93<br>1.000 | 1.12<br>1.000 | 0.50<br>1.000 | 0.11<br>1.000 | 0.40<br>1.000 |

## Discussion

COVID-19 confinement measures were associated with a clear decline in the number of patients presenting to the emergency services for i.e. heart problems, bowel obstruction and appendicitis [5]. Only sparse data are available on the relation of COVID-19 confinement on the number of emergency surgical procedures. This study investigated the influence of COVID-19 confinement measures on the number and composition of patient undergone appendectomy. The study revealed a significant decrease of appendectomies during the lockdown in 2020 compared to 2019 and 2018 in Germany.

Similar to our results, Tanel et al. reported data from Israel showing a decrease of acute appendicitis cases, which was not accompanied by an increase of complicated cases [6].

A single center study from New York, focusing on pediatric patients, showed no differences in the number of appendicitis cases during the lockdown [7]. However, this study revealed a high number of patients treated non-operatively for appendicitis. Patients treated non-operatively were not investigated in our study.

Another single center study from Madrid reported a higher proportion of appendectomies due to complex acute appendicitis during the lockdown [8]. Our study showed similar results when rating the data relatively only for 2020.

The numbers of appendectomies for complex acute appendicitis remained unchanged in our series, whereas the numbers for simple acute and non-acute appendicitis were significantly reduced. Interestingly, the number of non-acute appendicitis was reduced by more than 50%. The proportion of acute complex appendicitis raised relatively in the investigated time period in 2020.

Looking at the subgroups, the reduction of appendectomies in female patients, especially due to simple acute or non-acute appendicitis is remarkable. It has been shown previously that female patients are overrepresented in appendectomies in several age groups, i.e. until 18 years [9]

It could be speculated, that during COVID-19 lockdown patients with mild symptoms were not seeking medical care because of concern about acquiring COVID-19 infection. Here additional research is needed, as data from outpatient medical care was not available at the time of analysis.

Another possible explanation for the observed reduction of appendectomy cases is that the hospitals focused only on urgent cases in accordance with the lockdown recommendations. Further, the reduced number of simple acute appendicitis did not lead to an increase of complex acute appendicitis, which could have been expected during the lockdown. Our study could not confirm results of previous reports, which showed an increase of complicated appendicitis. This might be due to the fact that these previous reports focused on very small, regional patient groups.

## Limitations

The study has a number of limitations. It is based on secondary analysis of administrative claims data. Under- or overdocumentation of individual diagnoses cannot be ruled out. Furthermore, there are limitations with regard to external validity of the patient characteristics and reported incidences since the patient collective studied was composed exclusively of AOK-insured persons. Although the collective of AOK-insured persons accounts for more than one-third of hospital cases in Germany, there are certain differences versus the population of persons insured by other statutory sickness funds in terms of the age structure and comorbidity profile [10]. The reported case reduction is also influenced by changes of the AOK collective between the years. The number of insured persons slightly rose from 2019 to 2020, so that case reductions might be slightly underestimated. However, the differences between the years were less than 2% within each group.

## Conclusion

The COVID 19 lockdown resulted in a reduced number of appendectomies but not in an increased number of acute complicated appendicitis. The study provides no evidence that the confinement measures resulted in a deterioration of medical care for appendicitis.

## Data Availability

The authors confirm that the data utilized in this study cannot be made available in the manuscript, the supplemental files, or in a public repository due to German data protection laws (Bundesdatenschutzgesetz, BDSG). Therefore, they are stored on a secure drive in the AOK Research Institute (WIdO), to facilitate replication of the results. Generally, access to data of statutory health insurance funds for research purposes is possible only under the conditions defined in German Social Law (SGB V Paragraph 287). Requests for data access can be sent as a formal proposal specifying the recipient and purpose of the data transfer to the appropriate data protection agency. Access to the data used in this study can only be provided to external parties under the conditions of the cooperation contract of this research project and after written approval by the sickness fund. For assistance in obtaining access to the data, please contact.

## Compliance with Ethical Standards

### Funding

No funding was received.

### Conflict of interest

There are no conflicts of interest and no competing interests.

### Ethical statement

This article does not contain any studies with human participants performed by any of the authors.

### Data availability

The authors confirm that the data utilized in this study cannot be made available in the manuscript, the supplemental files, or in a public repository due to German data protection laws (‘Bundesdatenschutzgesetz’, BDSG). Therefore, they are stored on a secure drive in the AOK Research Institute (WIdO), to facilitate replication of the results. Generally, access to data of statutory health insurance funds for research purposes is possible only under the conditions defined in German Social Law (SGB V § 287). Requests for data access can be sent as a formal proposal specifying the recipient and purpose of the data transfer to the appropriate data protection agency. Access to the data used in this study can only be provided to external parties under the conditions of the cooperation contract of this research project and after written approval by the sickness fund. For assistance in obtaining access to the data, please contact.”

## Notes

### Competing Interest Statement

The authors have declared no competing interest.

### Author Declarations

The ethics committee of the University Hospital Frankfurt, Frankfurt, Germany, where the corresponding author is affiliated, confirmed that no ethical approval was needed for this retrospective study based on anonymized claims data. Confirmation was given by: Ethikkommission des Fachbereichs Medizin, Universitaetsklinikum der Goethe-Universitaet, Theodor-Stern-Kai 7, Haus 1, 2. OG, Zimmer 207-211, 60590 Frankfurt am Main,, https://www.kgu.de/index.php?id=735

## References

1. De Filippo O, D’Ascenzo F, Angelini F, et al (2020) Reduced Rate of Hospital Admissions for ACS during Covid-19 Outbreak in Northern Italy. N Engl J Med 383:88–89. https://doi.org/10.1056/NEJMc2009166

2. Dreifuss NH, Schlottmann F, Sadava EE, Rotholtz NA (2020) Acute appendicitis does not quarantine: surgical outcomes of laparoscopic appendectomy in COVID-19 times. Br J Surg. https://doi.org/10.1002/bjs.11806

3. Snapiri O, Rosenberg Danziger C, Krause I, et al (2020) Delayed diagnosis of paediatric appendicitis during the COVID’ pandemic. Acta Paediatr 109:1672–1676. https://doi.org/10.1111/apa.15376

4. Kölfel W (2020), RP online, https://rp-online.de/nrw/panorama/kinderaerzte-warnen-eltern-zoegern-arztbesuche-wegen-corona-hinaus_aid-50230583

5. Masroor S (2020) Collateral damage of COVID’ pandemic: Delayed medical care. J Card Surg 35:1345–1347. https://doi.org/10.1111/jocs.14638

6. Tankel J, Keinan A, Blich O, et al (2020) The Decreasing Incidence of Acute Appendicitis During COVID-19: A Retrospective Multi-centre Study. World J Surg 44:2458–2463. https://doi.org/10.1007/s00268-020-05599-8

7. Kvasnovsky CL, Shi Y, Rich BS, et al (2020) Limiting hospital resources for acute appendicitis in children: Lessons learned from the U.S. epicenter of the COVID-19 pandemic. J Pediatr Surg 6–10. https://doi.org/10.1016/j.jpedsurg.2020.06.024

8. Velayos M, Muñoz-Serrano Aj, Estefanía-Fernández K, et al (2020) Influencia de la pandemia por coronavirus 2 (SARS-Cov-2) en la apendicitis aguda. An Pediatría 93:118–122. https://doi.org/10.1016/j.anpedi.2020.04.022

9. Rolle U, Maneck M (2016) Versorgungstrends, regionale Variation und Qualität der Versorgung bei Appendektomien. In: Klauber J, Günster c, Schmacke N, et al (eds) Versorgungs report 2015/2016. Schattauer, Stuttgart, pp 217–38

10. Hoffmann F, Icks A (2012) Unterschiede in der Versichertenstruktur von Krankenkassen und deren Auswirkungen für die Versorgungsforschung: Ergebnisse des Bertelsmann-Gesundheitsmonitors. Das Gesundheitswes 74:291–297. https://doi.org/10.1055/s-0031-1275711

